## Supplementary Materials for "Different antigenic distance metrics generate similar predictions of influenza vaccine response breadth despite moderate correlation"

### 1. Reproducibility instructions

In order to reproduce our results you should first download the archived repository from either Zenodo (DOI: 10.5281/zenodo.15522148) or clone/download the Git repository (hosted on GitHub here: https://github.com/ahgroup/billings-comp-agdist-public). Note that if you use different software, software versions, or run the results in a way that differs from these instructions you may have expected errors or differences between your results and ours.

We ran our analysis pipeline on the University of Georgia's sapelo2 computing cluster, which is a distributed computing cluster running CentOS Linux release 7.5 which uses Slurm to schedule jobs. Our code is written as a `targets` pipeline (1) and can detect whether you are running an HPC job in a Slurm environment or not. Notably if you use a Slurm cluster computing environment which is configured differently from UGA's sapelo2 environment, you may need to make changes to the Slurm submission script (`job.sh`) or to the portion of the `_targets.R` script that defines the Slurm jobs. Our code will run on a local interactive R session as well, and will automatically detect the number of cores available to use. **Each Bayesian model currently requests 32 cores and we therefore highly suggest running the main analysis on a cluster computing setup.**

Once you've downloaded the code, you should open the `.Rproj` file in Rstudio. Using the R project file and RStudio is not mandatory, but if you don't, we assume you know what you are doing. You can then run the entire analysis pipeline by running `targets::tar_make()` in the console. If you are in an extremely limited computing environment, you can add the option `use_crew = FALSE` to force all targets to execute sequentially (although in this case you probably do not want to run the bayesian models anyways). You can run the command `targets::tar_visnetwork(TRUE)` to see an interactive graph of our analysis pipeline, and you can pass a vector of target names to `tar_make()` (potentially using `tidyselect` to define the vector) to only run those targets. Note that due to differences in OS and file systems, targets will likely appear outdated for you even though they are up-to-date. We also do not provide all of our model result files in the GitHub because they are extremely large (over 100GB) and infeasible to distribute, so if you want to edit or examine the Bayesian models you will need to rerun the code.

You will need the following software requirements to run our code.

- R version 4.4.1, available from https://cran.r-project.org.
- A working C++ compiler – on a linux cluster this is probably set up for you already, but on Windows you will need RTools 4.4, which is also available from CRAN. On MacOS you will need the latest version of the XCode command line tools.
- The RStudio IDE, available from https://posit.co/download/rstudio-desktop.
- Quarto version 16.40, available from https://quarto.org.

- The renv R package, version 1.1.4, available from https://cran.r-project.org/web/packages/renv/index.html. It will also attempt to install itself the first time you open our R project.
- Multiple system dependencies, including `CMake`. On Windows/MacOS these are provided by RTools or XCode respectively. On any type of linux there may be additional system requirements you will need to download. Your system should prompt you about this.
- The packages specified in the file `renv.lock`, which can be installed as explained in the next section.

With the software installed, follow these instructions to reproduce our results.

1. Open the `billings-comp-agdist-public.Rproj` file in Rstudio.
2. Once `renv` initializes, run the command `renv::restore()` in the console to begin installing the required packages. If you have issues at this stage you can also install the dependencies manually, but if you do not use `renv` or you use different package versions than we did, our code might not work for you.
3. If you want to run any steps that involve Bayesian models, you need to install `cmdstan` following the `cmdstanr` quick start guide at this location: https://mc-stan.org/cmdstanr/articles/cmdstanr.html. We used cmdstan version 2.36.0 for this project.
4. Now you can run our pipeline by running `targets::tar_make()` in the console. If you are new to targets and want to learn more about how the pipeline works, we recommend reading the targets manual which can be found here: https://books.ropensci.org/targets/.

**Again we note that our code is computationally intensive and we ran it on a distributed computing cluster. It still took multiple days to run, even running many operations in parallel with many cores each.**

### 2. Extended Methods

#### 2.1 Antigenic distance calculation

We calculated four different antigenic distance metrics for our study. In this section, we walk through how each method is calculated. Note that we only considered pairwise distances between strains of the same subtype. So we only computed distances between two A(H1N1) strains, between two A(H3N2) strains, or between two influenza B strains, we did not compute distances between A(H1N1) and A(H3N2) strains or between any A and B strains. However, since the two B lineages are quite similar and our panel included pre-divergence influenza B strains, we performed pairwise comparisons of all influenza B strains.

**Temporal distance** is the absolute value of the difference in the years of isolation between the two strains. For example, the difference between A/H1N1/California/09 and A/H1N1/Michigan/15 would be $|2015 - 2009| = 6$. Notably, in our study, we did not have any examples where the assay strain was isolated later than the vaccine strain, so taking the absolute value is not necessary, but we wanted to avoid confusion about our definitions. Future studies that collect such data might prefer either “backwards” or “forwards” temporal distances, and we can’t comment on that here.

**Dominant $p$-Epitope distance** is the maximum length-normalized Hamming distance across the five major epitope sites on the HA head. After aligning the HA amino acid sequences for all of the strains, we removed the signal peptides from the sequences and used the previously identified epitope site locations for influenza A (2) and influenza B (3). Working pairwise with the sequences, we concatenated the residues for each epitope and calculated the Hamming distance between each epitope, and we divided the Hamming distance for a given epitope by the number of residues in that epitope. Then the $p$-Epitope distance for that pair of strains was the maximum of those epitope-wise distances.

**Grantham’s distance** is a weighted distance based on biochemical properties that considers how different two differing residues at the same position are. We used Grantham’s substitution matrix (4) to assign a value to each residue site between two sequences, based on the transition between amino acids. More different transitions are given higher weights. Then, for each pair of sequences, we sum the weights for that pair and divide by the length of the sequence.

Finally, **cartographic distance** is the Euclidean distance between strains on antigenic cartography map. We built our cartographic maps from the combined table of post-vaccination titer data in our study, treating all person-years as independent occurrences (there is no clear meaning for repeat measurements in a dimension reduction analysis). We used `Racmacs`, which implements metric multidimensional scaling, to create and optimize the cartographic map (5). All of our maps were two dimensional, and we selected the best fitting map from 25 distinct `Racmacs` runs with random initializations, where each initialization was allowed to perform up to 100 L-BFGS optimization runs to relax the initial

MDS cartography. Multiple optimization runs are necessary because different initial conditions can lead to different maps (6). Combining multiple runs by applying a method like generalized Procrustes analysis is theoretically possible (simple averaging won't work because rotation and scaling need to be taken into account) but has not yet been studied or published so we instead chose the one overall best run. We did not perform dimensional analysis to choose 2D maps, we chose them for ease of interpretation and based on previous convention.

For our models, we only considered the antigenic distance between the assay strain and the vaccine strain of the same subtype for a given HAI assay. Some of the assay strains used were influenza B strains isolated before the Victoria/Yamagata lineage divergence. Because our main question was about the antigenic distance, we compared pre-divergence B strains to both the Yamagata and Victoria vaccine strains in our analyses. To facilitate fair comparisons across subtypes and antigenic distance metrics, we min-max normalized the antigenic distance measurements within each combination of influenza season, subtype, and metric. After normalization, the antigenic distance for homologous measurements was set to 0, and the antigenic distance for the most different assay strain used in a given season was set to 1, with all other antigenic distance values falling in this interval.

#### 2.2 Sequence data sources

We retrieved HA sequences for each strain from either the U.S. National Center for Biotechnology Information (NCBI)'s GenBank database (7,8), the UniProt dataset (9), or GISAID's EpiFlu database (10,11). The attribution and accession numbers for each strain are listed in Table 1.

Most of the sequences we used from GenBank and UniProt are not associated with particular publications and are only able to be referenced via their accession numbers. The following sequences have formal references: `AAD17229.1` (12); `AAA67338.1` (13); `AAP34324.1` (14); `ADE28750.1` (15); `ACP41953.1` (16); `ABQ97200.1` (17); `AAA62338.1` (18); `AIW60702.1` (19); `P03460` and `P03461` (20); and `P12443` (21).

The sequences we used from GISAID are accessible via GISAID Identifier EPI_SET_250609vz and DOI https://doi.org/10.55876/gis8.250609vz. To view the contributors of each individual sequence with details such as accession number, Virus name, Collection date, Originating Lab and Submitting Lab and the list of Authors, visit 10.55876/gis8.250609vz.

*Table 1: Accession number and source for each HA sequence used in our analysis.*

| Strain Name | Source | Accession # |
|---|---|---|
| A/H1N1/South Carolina/1/1918 | GenBank | AAD17229.1 |
| A/H1N1/Puerto Rico/8/1934 | GenBank | AGU93019.1 |
| A/H1N1/Weiss/1943 | GenBank | ABD79101.1 |
| A/H1N1/Fort Monmouth/1/1947 | GenBank | AAA67338.1 |
| A/H1N1/Denver/1957 | GenBank | ABD15258.1 |
| A/H1N1/New Jersey/8/1976 | GenBank | AGB51356.1 |
| A/H1N1/Ussr/90/1977 | GenBank | ABD95350.1 |

| | | |
|---|---|---|
| A/H1N1/Brazil/11/1978 | GenBank | ABO38065.1 |
| A/H1N1/California/10/1978 | GenBank | ABP49338.1 |
| A/H1N1/Chile/1/1983 | GenBank | ABO38340.1 |
| A/H1N1/Singapore/6/1986 | GenBank | ABO38395.1 |
| A/H1N1/Texas/36/1991 | GenBank | ACF41933.1 |
| A/H1N1/Beijing/262/1995 | GenBank | ACF41867.1 |
| A/H1N1/New Caledonia/20/1999 | GenBank | AAP34324.1 |
| A/H1N1/Solomon Islands/3/2006 | GenBank | ABU99109.1 |
| A/H1N1/Brisbane/59/2007 | GenBank | ADE28750.1 |
| A/H1N1/California/07/2009 | GenBank | ACP41953.1 |
| A/H1N1/Michigan 45/2015 | GenBank | AMV49034.1 |
| A/H1N1/Brisbane/02/2018 | GISAID | EPI1415369 |
| A/H1N1/Guangdong-Maonan/SWL1536/2019 | GISAID | EPI3133357 |
| A/H1N1/Victoria/2570/2019 | GenBank | WEY08940.1 |
| A/H3N2/Hong Kong/8/1968 | GenBank | ABQ97200.1 |
| A/H3N2/Port Chalmers/1/1973 | GenBank | ABE12532.1 |
| A/H3N2/Texas/1/1977 | GenBank | AFM68965.1 |
| A/H3N2/Mississippi/1/1985 | GenBank | AAA62338.1 |
| A/H3N2/Sichuan/2/1987 | GenBank | AFG72085.1 |
| A/H3N2/Shandong/9/1993 | GenBank | AFH00285.1 |
| A/H3N2/Nanchang/933/1995 | GenBank | AFG72625.1 |
| A/H3N2/Sydney/5/1997 | GenBank | ACO95259.1 |
| A/H3N2/Panama/2007/1999 | GenBank | ABF21273.1 |
| A/H3N2/Fujian/411/2002 | GenBank | AFG72823.1 |
| A/H3N2/New York/55/2004 | GenBank | ACF41900.1 |
| A/H3N2/Wisconsin/67/2005 | GenBank | AHG96791.1 |
| A/H3N2/Brisbane/10/2007 | GenBank | AIW60702.1 |
| A/H3N2/Uruguay/716/2007 | GenBank | ACD47213.1 |
| A/H3N2/Perth/16/2009 | GenBank | ACS71642.1 |
| A/H3N2/Victoria/361/2011 | GenBank | AIU46088.1 |
| A/H3N2/Texas/50/2012 | GenBank | AGL07159.1 |
| A/H3N2/Switzerland/9715293/2013 | GISAID | EPI530687 |
| A/H3N2/Hong Kong/4801/2014 | GISAID | EPI834581 |
| A/H3N2/Singapore/infimh-16-0019/2016 | GISAID | EPI780183 |
| A/H3N2/Kansas/14/2017 | GenBank | AVG71503.1 |
| A/H3N2/South Australia/34/2019 | GISAID | EPI1387331 |
| A/H3N2/Hong Kong/2671/2019 | GenBank | WMW30924.1 |
| A/H3N2/Tasmania/503/2020 | GenBank | WMW30850.1 |
| A/H3N2/Darwin/9/2021 | GenBank | WND60806.1 |
| B/Lee/1940 | UniProt | P03460 |
| B/Maryland/1959 | UniProt | P03461 |
| B/Singapore/1964 | UniProt | P12443 |
| B/Victoria/02/1987 | UniProt | A4D5N9 |
| B/Hong Kong/330/2001 | GenBank | ABL77178.1 |
| B/Malaysia/27127/2004 | GenBank | AFJ80733.1 |
| B/Victoria/326/2006 | GenBank | AGX18732.1 |
| B/Brisbane/60/2008 | GenBank | AFH57909.1 |
| B/Colorado/06/2017 | GenBank | ASK81305.1 |
| B/Washington/02/2019 | GenBank | WIM08940.1 |
| B/Michigan/01/2021 | GenBank | WMW30908.1 |
| B/Austria/1359417/2021 | GISAID | EPI1868375 |
| B/Yamagata/16/1988 | GenBank | ABL77255.1 |
| B/Harbin/7/1994 | GenBank | ACR15721.1 |
| B/Sichuan/379/1999 | GISAID | EPI2085837 |
| B/Florida/4/2006 | GenBank | ACA33493.1 |
| B/Wisconsin/01/2010 | GenBank | AET22057.1 |
| B/Texas/06/2011 | GenBank | AGI64713.1 |
| B/Massachusetts/02/2012 | GenBank | AGL06036.1 |
| B/Phuket/3073/2013 | GISAID | EPI3555941 |

#### 2.3 Causal modeling and model formulation

While we do not claim that our estimates are causal, we employed a graphical causal model to formulate our statistical models. While all statistical models are a mix between practicality and the best possible model, we hope that by formalizing our thinking, our models will be robust and correctly answer our research questions.

Our original dataset contained one record per HAI assay, indicating the individual, season, study site, time point (pre- or post-vaccination), vaccine dose, and assay strain for each record. The data also included the following demographic variables: age, birth year, sex assigned at birth, and reported race/ethnicity. The study also provided a list of vaccine strains for each formulation (see the section on vaccine formulation for a complete list). Note that we only analyzed standard dose vaccine recipients in our analysis, so we do not discuss the vaccine dose further.

We built a causal model for the effect of antigenic distance as a directed acyclic graph (DAG). We include the following variables in our causal model: $U$, unobserved confounders that could be partially explained by nuisance variation, but are not directly explained in our model; $p$, the pre-vaccination titer; $C$, the set of individual covariates which could potentially impact both pre-vaccination and post-vaccination titers; $sv$, the vaccine strain (for a given subtype); and $sa$, the assay strain for a particular HAI assay. We considered $C$ to represent all observed demographic features reported in our study, including age, birth year, sex assigned at birth, and race/ethnicity. Other sources of nuisance variation, including the study site an individual reported to and other sources of individual variation which we have not observed, are encoded in the unobserved confounder $U$. The causal model we selected is shown in Figure 1.

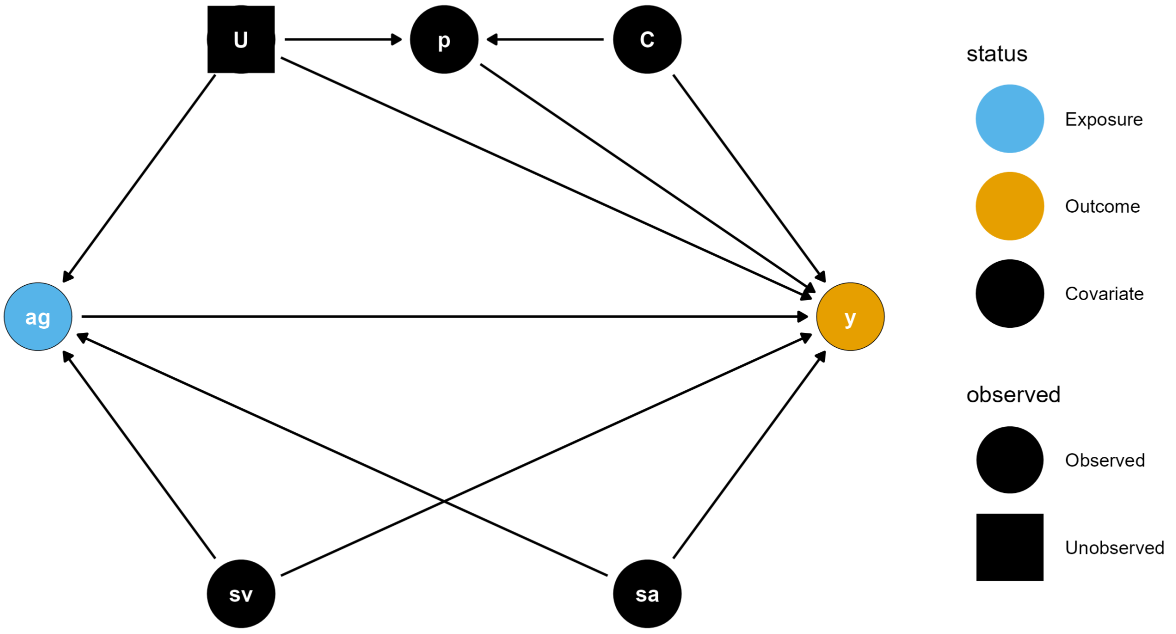

*Figure 1: The graphical causal model for our research question represented as a DAG.*

Under this causal model, the only confounders are the vaccine strain and assay strain, and any unobserved confounders. If we assume no unmeasured confounding, then the minimal sufficient adjustment set is only the vaccine strain and assay strain. However, our goal in this project was to analyze the effect of antigenic distance as a predictor without incorporating strain-specific effects. So, we stratified our models by vaccine strain (i.e., we fit all models separately for each vaccine component) and deliberately did not include a strain-specific effect.

To adjust for nuisance variation (potentially a source of unmeasured confounding), we included random effects to control for measurements at the same study site and on the same individual. Finally, we included specific ancestors of the outcome variable, which is not necessary to close backdoor paths and does not mitigate any biases in our estimates. However, including these variables can improve the efficiency of the estimators of interest. We included pre-vaccination titer and age specifically in our model. In our previous work, we found that sex and race/ethnicity have minimal association with the observed HAI titers, but we included them in our model for completeness. Since the majority of participants in our study were white (see the demographics table in a later section), we coded race/ethnicity as an indicator variable that was equal to 0 if the participant identified as white or Caucasian and not Hispanic or Latino, and 1 otherwise. We coded sex as 0 if the participant's sex assigned at birth was reported as male and 1 if it was reported as female. We included pre-vaccination titer in the model as-is, but since the age has a large range (from 11 to 65), we minmax scaled the age before using it in a model. Minmax scaling variables with large ranges can improve numerical stability of the model, but the model can still make predictions for any age. Similarly, we minmax scaled the birth year.

Finally, we note that in some models it is also possible for cross-season differences to exist when the same vaccine strain was used for multiple years in a row. I.e., we might expect post-vaccination titers to change due to repeated usage of the same vaccine. However, since some of the vaccine strains were only used for one year before being replaced, this seasonal effect is not estimable in all of our models. Therefore, we decided not to include a seasonal effect in any of the models, especially since the effect of repeated usage of the same vaccine strain was not our primary research question.

#### 2.4 Model implementation

We fit two models using `brms`, a generalized additive mixed model (GAMM) and a linear mixed model (LMM). The models were identical other than the specification for the effect of antigenic distance, so we will first describe the general parts of the model. Note that in the following mathematical descriptions, we adopt bracket notation rather than subscript notation following the convention of McElreath (22) due to the large number of subscripts in our model. That is, we use the notation $y[i]$ in place of the conventional $y_i$. We use subscripts to instead identify unique parameters. We also used the centered dot symbol $(\cdot)$ to avoid repetition when there are many valid arguments that would have the same right-hand side in a formula. For example, $\zeta[\cdot]$ indicates that all subscripts for $\zeta$ use the same equation.

We modeled our outcome (post-vaccination titer) as a Gaussian random variable, but due to the censored nature of our data we applied a censoring correction in the likelihood. Letting the outcome for a specific vaccine component be $y$, we assumed that

$$
\begin{aligned}
f(y[i] \mid \mu[i], \sigma^2) &= \int_{L[i]}^{U[i]} \mathcal{N}\,(y[i] \mid \mu[i], \sigma^2)\ \mathrm{d}y[i] \\
\sigma &\sim t^{+}(3,0,1) \\
i &= 1, \dots, n
\end{aligned}
$$

where $L[i]$ and $U[i]$ are the lower and upper censoring bounds respectively (see the section on censoring bounds for details), $\mathcal{N}(\mu, \sigma^2)$ is the Gaussian (Normal) probability density function with mean $\mu$ and variance $\sigma^2$, $t^{+}(\nu, \mu, \sigma)$ is the location-scale half Student's $t$ distribution with degrees of freedom $\nu$, location parameter $\mu$, and scale parameter $\sigma$. We chose a Student's $t$ prior with $\nu = 3$ degrees of freedom because the distribution has fat tails, which allows the variance to be large if supported by the data, but we assume *a priori* that the distribution of the variance has a finite location and scale parameter (which is only the case when $\nu > 2$). Here, $i$ is the index for the current data record, representing one HAI assay, and $n$ is the total number of HAI assays (records) in the dataset.

The model for the mean is shown below, including the priors for each parameter. For now, we represent the effect of antigenic distance as a function $g$, which we detail with its priors in the next section. (Note that here we refer to the $\beta_j$ with subscripts because we treat each as an independent parameter rather than as a vector of parameters, which would be implied by $\beta[j]$. There is conceptually little difference between these notational approaches but we feel that this notation better emphasizes the independent priors on each $\beta_j$.)

$$
\begin{aligned}
\mu[i] &= \beta_0 + u\big[1,\text{id}[i]\big] + u\big[2,\text{study}[i]\big] + u\big[3,\text{subtype}[i]\big] + \\
&\quad u\big[4,\text{subtype}[i] \times \text{vaccine strain}[i]\big] + u\big[5,\text{subtype}[i] \times \text{assay strain}[i]\big] + \\
&\quad g(\text{antigenic distance}[i]) + \\
&\quad \beta_p(\text{log pre-vaccination titer[i]}) + \beta_a(\text{scaled age}[i]) + \\
&\quad \beta_y(\text{scaled birth year}[i]) + \beta_r(\text{race/ethnicity[i]}) + \beta_s(\text{sex}[i]) \\
\beta_{(\cdot)} &\sim \mathcal{N}(0,1) \\
u[r,\cdot] &\sim \mathcal{N}(0, \omega[r]) \quad r = 1,2,\dots,5 \\
\omega[r] &\sim t^{+}(3,0,1)
\end{aligned}
$$

The priors follow the same formulation as before, but we chose Gaussian priors for the beta effects. Gaussian priors have flatter tails than Student's $t$ priors, which provides a more regularizing effect for the beta parameters – that is, we presuppose that they are more likely to be close to zero, and our data needs to be strong enough to move the posterior distributions away from zero before we can make any conclusions.

The functional form of $g$ is the only difference between the GAMM and the LMM. In the LMM, $g$ takes a simple linear form:

$$
\begin{aligned}
g(\text{antigenic distance}[i]) &= \beta_d(\text{antigenic distance}[i]) \\
\beta_d &\sim \mathcal{N}(0,1)
\end{aligned}
$$

where the antigenic distance is minmax normalized for each model as described in the antigenic distance calculation section. For the GAMM, the function form of $g$ is more complex. We modeled the antigenic distance effect using a thin-plate basis spline, which allows for the relationship to be curved in an arbitrary pattern, but constrains the fit so that rapid changes in the pattern are penalized and must be supported by data (23–27). The specific form is

$$
\begin{aligned}
g(\cdot) &= \sum_{k=1}^{5} \gamma\,[k] \cdot \phi[k](\cdot) \\
\gamma[k] &\sim \mathcal{N}(0, \tau) \\
\tau &\sim t^{+}(3{,}0{,}0.25)
\end{aligned}
$$

where the $\gamma[k]$ are coefficients which are regularized to be similar via an adaptive prior and the $\phi[k]$ are thin-plate spline basis functions. Thin-plate splines use a low-rank approximation of the spline basis for computational efficiency, which can be tuned to balance between accuracy and efficiency. The maximal $k$ (or size of the spline basis) we can choose is equal to the number of unique values of the predictor, so we chose $k = 5$, which was estimable across all of our antigenic distance metrics. We used Student's $t$ priors for the adaptive prior on the variance of the spline coefficients so that the spline can be wiggly if supported by the data, but we chose a conservative hyperprior variance (0.25, based on a prior predictive simulation) to constrain the spline towards being flat if the signal from the data is not strong.

The random effects we included in the model represented sources of nuisance variation which we were interested in controlling for, but not specifically estimating. We included random effects to capture interindividual variation, variation across study sites, and direct effects of the influenza strains not explained by the antigenic distance. We included random intercepts for individuals ($u[1,\cdot]$) and study sites ($u[2,\cdot]$) in a typical way with regularizing priors. To control for the direct effects of influenza assay and vaccine strains, we noted that each strain was nested within a subtype, but the assay strains and vaccine strains were not themselves crossed or nested (each assay strain could appear with an arbitrary combination of different vaccine strains, although all assay strains and all vaccine strains are only ever associated with a single influenza subtype). Including the subtype effect as $u[3,\cdot]$ and then including random effects which consider both the subtype and the vaccine strain ($u[4,\cdot]$) or the assay strain ($u[5,\cdot]$) allows for assay/vaccine strains within the same subtype to have a correlated effect, while assay/vaccine strains of different subtypes do not have correlated effects. Again, we assigne skeptical, regularizing priors to all of these random effects.

#### 2.5 Censoring bounds

HAI titer assays, like all serial dilution assays, produce censored data values. In fact, all values produced by an HAI assay are censored. We take this censoring into account in the likelihood of our model by integrating over the censoring bounds for a given data point $y_i$.

All serial dilution assays are censored – for the case of HAI, we assume that there is some latent, true dilution $y_i^*$ which is the minimal dilution where hemagglutination is not observed. This is likely some decimal number, and we will never observe this true value. Instead, we chose a starting dilution, $y_{\min}$, which is 10 in our dataset. If we observe agglutination at this starting dilution, we say the value is below the limit of detection and it is recorded as 5 in our dataset. These values are left censored. In reality, we know that the latent agglutination dilution for an assay can be any value less than 10, i.e., our censoring bounds for these assays are $(0,10)$.

There is also a maximal dilution for the assay, $y_{\max}$, which was 20480 in our dataset. In practice, if researchers don't observe hemagglutination at any dilution, they can simply continue diluting the assay until they observe agglutination. However, a standard 96-well plate only has 12 columns, so most studies will report 20480 (the 12th serial dilution for an HAI assay starting at 10 and doubling each dilution). So these values are right censored, and the censoring bounds are $[20480, \infty)$. Note that the lower bound of the interval is included because the value *could* be exactly 20480 (though this occurs with probability zero for a continuous latent variable).

Finally, any other assay with a result between the limits of detection will also be interval censored, because we only observe certain dilutions. For example, if we observe inhibited hemagglutination at a dilution of 10, but agglutination occurs at a dilution of 20, we record the result as 10. However, we don't know that a dilution of 1:15 wouldn't cause inhibition, so we only know that the latent dilution is in the interval $[10,20)$. Similarly for any value $y_{\min} < y < y_{\max}$, the latent dilution is in the interval $[y, 2y)$.

Converting to the log scale, the censoring bounds $L$ and $U$ that we refer to in the previous equations are as follows (here we omit subscripts to avoid confusion with interval notation, but $L$, $U$, and $y$ all vary by individual while $y_{\min}$ and $y_{\max}$ are constant):

$$(L, U) = \begin{cases} (-\infty, y_{\min}) & y = y_{\min} \\ [y, y+1), & y_{\min} < y < y_{\max} \\ [y_{\max}, \infty) & y = y_{\max} \end{cases}.$$

For our study, $y_{\min} = \log_2(10/5) = 1$ and $y_{\max} = \log_2(20480/5) = 12$.

#### 2.6 Stan implementation

We obtained posterior samples of the model parameters using the No U-Turn Sampler (NUTS) algorithm implemented by Stan (28,29), via the `brms` (30–32) and `cmdstanr` (33) packages for `R` (34). In `brms`, we specified our model formulas as

```
y | cens(c, y2) ~ 1 +
    birth_year_c + age_c + sex_i + race_i +
    log_pretiter + s(d_norm, k = 5, by = strain_type) +
    (1 | strain_type) +
    (1 | study) + (1 | subject_id) +
    (1 | strain_type:vaccine_name) + (1 | strain_type:strain_name)
```

for the GAMMs and

```
y | cens(c, y2) ~ 1 +
    birth_year_c + age_c + sex_i + race_i +
    log_pretiter + d_norm + (1 + d_norm | strain_type) +
    (1 | study) + (1 | subject_id) +
    (1 | strain_type:vaccine_name) + (1 | strain_type:strain_name)
```

for the LMMs. We specified our prior distributions as

```
brms::prior(normal(0,1), class = "Intercept"),
brms::prior(normal(0,1), class = "b"),
brms::prior(student_t(3, 0, 1), class = "sd", lb = 0),
brms::prior(student_t(3, 0, 1), class = "sigma", lb = 0),
brms::prior(student_t(3, 0, 0.25), class = "sds", lb = 0)
```

for the GAMMS and

```
brms::prior(normal(0,1), class = "Intercept"),
brms::prior(normal(0,1), class = "b"),
brms::prior(student_t(3, 0, 1), class = "sd", lb = 0),
brms::prior(student_t(3, 0, 1), class = "sigma", lb = 0)
```

for the LMMs. Note that the LMMs do not have a prior for parameters of class `sds`, which represent the adaptive smoothing priors for spline coefficients. We sampled the models on 32 chains with 200 warmup iterations and 625 post-warmup sampling iterations per chain for a total of 20,000 posterior post-warmup samples for each parameter. The effective number of samples is shown in the model diagnostics table in a later section. We also specified an adaptive delta of 0.99 and a maximum treedepth of 12, and used the recommended Stan and `brms` default values for all other algorithm control samples. Notably, this means each chain was initialized with a random variable – choosing smart initial values could potentially speed up the sampling, but we found that this was not sufficient to warrant further investigation for our models. The primary cause of slow sampling for our models was the large number of data points, although we also have several hierarchical parameters which can slow sampling.

#### 2.7 Posterior marginal effects

To summarize our models, in Figure 2 we present a posterior contrast that we call a marginal effect, but is technically a marginal conditional effect that marginalizes some variables and is conditional on others. The posterior effect of interest is the effect of antigenic distance on post-vaccination titer, conditional on the subtype for each antigenic distance metrics (note that models were fit completely separate for each antigenic distance metric). In order to calculate this effect, we constructed counterfactual predictions to estimate for our model on an interpolated grid of antigenic distance values using the `marginaleffects` package (35).

The effects we present in main text Figure 2 (for each separate antigenic distance metric, which were all fit as completely separate models with the same set of effects other than the different antigenic distances) are conditional on the subtype, and represent global means of the marginal effects at the mean for the other variables included. This means that are results are conditional on the random effects in the model, but do not include random effects variances in the credible intervals for effects other than the subtype.

We specified interpolated counterfactual values of the normalized antigenic distance from 0 to 1, spaced by 0.01, excluded the effects of random effects parameters, set categorical fixed effects values to their mode, and set continuous fixed effects (other than antigenic distance) to their mean. In this way, our marginal effects represent the expected post-vaccination titer for this population for a typical individual who is similar to those included in our study population. While using average marginal effects (AMEs) allows us to generalize our predictions to other levels of the fixed effects for a typical individual, and we could integrate out the random effects, we found that this more computationally demanding methods only served to inflate the credible interval without substantively changing the predictions we made. Since the credible interval is already quite wide, and should be interpreted conservatively as in all non-causal observational studies, we did not see the need to include nuisance variation in our predictions, since the main focus of our study was comparing the similarity of predictions across the four antigenic distance metric models, rather than specifically trying to isolate a causal effect of antigenic distance.

Specifically, we obtained predictions from the models on a grid defined by the following `marginaleffects` package syntax.

```
marginaleffects::datagrid(
            model = model_i,
            d_norm = seq(0, 1, 0.01),
            strain_type = c("H1N1", "H3N2", "B-Vic", "B-Yam")
        )
```

Here, `model_i` refers to each of the models that we fit.

### 3. Supplementary results

#### 3.1 Annual Fluzone vaccine formulation

Table 2 shows the strains which were included in each season's formulation of the Fluzone vaccine. We only show the formulation for the standard dose (SD) vaccine (the HD vaccine was trivalent throughout the study years we selected, while the quadrivalent formulation of the SD vaccine became available in 2015/16).

*Table 2: Strains used in the Fluzone standard dose vaccine formulation during each influenza season.*

| Season | A(H1N1) | A(H3N2) | B/Victoria | B/Yamagata |
|---|---|---|---|---|
| 2013/14 | CA/09 | TX/12 | — | MA/12 |
| 2014/15 | CA/09 | TX/12 | — | MA/12 |
| 2015/16 | CA/09 | Switz/13 | Bris/08 | Phu/13 |
| 2016/17 | CA/09 | HK/14 | Bris/08 | Phu/13 |
| 2017/18 | MI/15 | HK/14 | Bris/08 | Phu/13 |

#### 3.2 Annual heterologous strain panel

The strains used in each panel are shown in Table 3. A shaded cell with an X in it indicates that the strain indicated by the current row was used as part of the HAI panel in the season indicated by the current column.

*Table 3: Heterologous strain panel used during each influenza season.*

| Subtype | Strain | 2013/14 | 2014/15 | 2015/16 | 2016/17 | 2017/18 |
|---|---|---|---|---|---|---|
| A(H1N1) | SC/18 | X | X | X | X | X |
| | PR/34 | X | | | | |
| | Wei/43 | X | X | X | X | X |
| | FM/47 | X | X | X | X | X |
| | Den/57 | X | X | X | X | X |
| | NJ/76 | X | X | X | X | X |
| | USSR/77 | X | X | X | X | X |
| | Bra/78 | X | | | X | X |
| | CA/78 | | X | X | | |
| | Chi/83 | X | X | X | X | X |
| | Sing/86 | X | X | X | X | X |
| | TX/91 | X | X | X | X | X |
| | Bei/95 | X | X | X | X | X |
| | NC/99 | X | X | X | X | X |
| | SI/06 | X | X | X | X | X |

| | | | | | | |
|---|---|---|---|---|---|---|
| | Bris/07 | X | X | X | X | X |
| | CA/09 | X | X | X | X | X |
| | MI/15 | | | | X | X |
| A(H3N2) | HK/68 | X | X | X | X | X |
| | PC/73 | X | X | X | X | X |
| | TX/77 | X | X | X | X | X |
| | MI/85 | X | X | X | X | X |
| | Sich/87 | X | X | X | X | X |
| | Shan/93 | X | X | X | X | X |
| | Nan/95 | X | X | X | X | X |
| | Syd/97 | X | X | X | X | X |
| | Pan/99 | X | X | X | X | X |
| | Fuj/02 | X | X | X | | |
| | NY/04 | X | X | X | X | X |
| | Br/07 | X | | | | |
| | WI/05 | X | X | X | X | X |
| | Uru/07 | | X | X | X | X |
| | Per/09 | X | X | X | X | X |
| | Vic/11 | X | X | X | X | X |
| | TX/12 | X | X | X | X | X |
| | Switz/13 | X | X | X | X | X |
| | HK/14 | | X | X | X | X |
| | Sing/16 | | | | | X |
| B/Pre | Lee/40 | X | X | X | X | |
| | MD/59 | | X | X | X | |
| | Sing/64 | | X | X | X | |
| B/Victoria | Vic/87 | | | | X | X |

| | | | | | | |
|---|---|---|---|---|---|---|
| | HK/01 | | | X | X | X |
| | Mal/04 | | | X | X | X |
| | Vic/06 | | | X | X | X |
| | Bris/08 | | | X | X | X |
| | CO/17 | | | X | X | X |
| B/Yamagata | Yam/88 | X | X | X | X | X |
| | Harb/94 | X | X | X | X | X |
| | Sich/99 | X | X | X | X | X |
| | FL/06 | X | X | X | X | X |
| | WI/10 | X | X | X | X | X |
| | TX/11 | X | X | X | X | X |
| | MA/12 | X | X | X | X | X |
| | Phu/13 | X | X | X | X | X |

#### 3.3 Strain names and abbreviations

Throughout the manuscript, we use abbreviated names for each strain. Table 4 shows the corresponding abbreviation for the full name of each strain.

*Table 4: Full strain names and associated abbreviations for each strain used in the study.*

| Subtype | Strain name | Short name |
|---|---|---|
| A(H1N1) | A/H1N1/South Carolina/1/1918 | SC/18 |
| | A/H1N1/Puerto Rico/8/1934 | PR/34 |
| | A/H1N1/Weiss/1943 | Wei/43 |
| | A/H1N1/Fort Monmouth/1/1947 | FM/47 |
| | A/H1N1/Denver/1957 | Den/57 |
| | A/H1N1/New Jersey/8/1976 | NJ/76 |
| | A/H1N1/Ussr/90/1977 | USSR/77 |
| | A/H1N1/Brazil/11/1978 | Bra/78 |
| | A/H1N1/California/10/1978 | CA/78 |
| | A/H1N1/Chile/1/1983 | Chi/83 |
| | A/H1N1/Singapore/6/1986 | Sing/86 |
| | A/H1N1/Texas/36/1991 | TX/91 |
| | A/H1N1/Beijing/262/1995 | Bei/95 |
| | A/H1N1/New Caledonia/20/1999 | NC/99 |
| | A/H1N1/Solomon Islands/3/2006 | SI/06 |
| | A/H1N1/Brisbane/59/2007 | Bris/07 |

| | | |
|---|---|---|
| | A/H1N1/California/07/2009 | CA/09 |
| | A/H1N1/Michigan 45/2015 | MI/15 |
| A(H3N2) | A/H3N2/Hong Kong/8/1968 | HK/68 |
| | A/H3N2/Port Chalmers/1/1973 | PC/73 |
| | A/H3N2/Texas/1/1977 | TX/77 |
| | A/H3N2/Mississippi/1/1985 | MI/85 |
| | A/H3N2/Sichuan/2/1987 | Sich/87 |
| | A/H3N2/Shandong/9/1993 | Shan/93 |
| | A/H3N2/Nanchang/933/1995 | Nan/95 |
| | A/H3N2/Sydney/5/1997 | Syd/97 |
| | A/H3N2/Panama/2007/1999 | Pan/99 |
| | A/H3N2/Fujian/411/2002 | Fuj/02 |
| | A/H3N2/New York/55/2004 | NY/04 |
| | A/H3N2/Brisbane/10/2007 | Br/07 |
| | A/H3N2/Wisconsin/67/2005 | WI/05 |
| | A/H3N2/Uruguay/716/2007 | Uru/07 |
| | A/H3N2/Perth/16/2009 | Per/09 |
| | A/H3N2/Victoria/361/2011 | Vic/11 |
| | A/H3N2/Texas/50/2012 | TX/12 |
| | A/H3N2/Switzerland/9715293/2013 | Switz/13 |
| | A/H3N2/Hong Kong/4801/2014 | HK/14 |
| | A/H3N2/Singapore/infimh-16-0019/2016 | Sing/16 |
| B/Pre | B/Lee/1940 | Lee/40 |
| | B/Maryland/1959 | MD/59 |
| | B/Singapore/1964 | Sing/64 |
| B/Victoria | B/Victoria/02/1987 | Vic/87 |
| | B/Hong Kong/330/2001 | HK/01 |
| | B/Malaysia/27127/2004 | Mal/04 |
| | B/Victoria/326/2006 | Vic/06 |
| | B/Brisbane/60/2008 | Bris/08 |
| | B/Colorado/06/2017 | CO/17 |
| B/Yamagata | B/Yamagata/16/1988 | Yam/88 |
| | B/Harbin/7/1994 | Harb/94 |
| | B/Sichuan/379/1999 | Sich/99 |
| | B/Florida/4/2006 | FL/06 |
| | B/Wisconsin/01/2010 | WI/10 |
| | B/Texas/06/2011 | TX/11 |
| | B/Massachusetts/02/2012 | MA/12 |
| | B/Phuket/3073/2013 | Phu/13 |

#### 3.4 Demographic information

A summary of the demographic information for the individuals included in our analysis is shown in Table 5, and includes information about their reported race/ethnicity, sex assigned at birth, age at first enrollment, and year of birth. The majority of participants identified their race as White or Caucasian, and were assigned female at birth. All participants from the PA and FL study sites were adults, but the UGA study site also recruited teenagers, and all three study sites included elderly people over 65 years of age. Most participants returned to the study site in at least one subsequent year, contributing more than one person-year of data to the study.

*Table 5: Demographic characteristics of the study participants. Summary statistics shown are count and column percent for sex, race, and contributed person-years; and median with range for age at first enrollment, birth year, and contributed HAI assays. Demographic variables were collected by a questionnaire from participants on the date they enrolled in a study season and received a vaccine. Coding details for the demographic variables are in the Supplement.*

| **Characteristic** | **FL**<br>N = 241 | **PA**<br>N = 133 | **UGA**<br>N = 303 | **Overall**<br>N = 677 |
|---|---|---|---|---|
| Sex Assigned at Birth, n (%) | | | | |
| Female | 184 (76) | 93 (70) | 168 (55) | 445 (66) |
| Male | 57 (24) | 40 (30) | 135 (45) | 232 (34) |
| Race/Ethnicity, n (%) | | | | |
| White | 190 (79) | 70 (53) | 233 (77) | 493 (73) |
| Black or African American | 14 (6) | 52 (39) | 24 (8) | 90 (13) |
| Other | 12 (5) | 8 (6) | 33 (11) | 53 (8) |
| Hispanic or Latino | 24 (10) | 3 (2) | 13 (4) | 40 (6) |
| Unknown | 1 (0) | 0 (0) | 0 (0) | 1 (0) |
| Age at First Enrollment, Median (Min - Max) | 42 (20 - 80) | 60 (26 - 81) | 25 (12 - 83) | 40 (12 - 83) |
| Year of Birth, Median (Min - Max) | 1972 (1933 - 1996) | 1954 (1932 - 1987) | 1991 (1934 - 2006) | 1975 (1932 - 2006) |
| Contributed HAI assays, Median (Min - Max) | 85 (40 - 189) | 94 (8 - 185) | 48 (47 - 95) | 52 (8 - 189) |
| Contributed person-years, n (%) | | | | |
| 1 | 114 (47) | 44 (33) | 206 (68) | 364 (54) |
| 2 | 52 (22) | 31 (23) | 97 (32) | 180 (27) |
| 3 | 61 (25) | 32 (24) | 0 (0) | 93 (14) |
| 4 | 14 (6) | 26 (20) | 0 (0) | 40 (6) |

Figure 2 shows a visualization of the collected pre-vaccination titers, and Figure 3 shows a visualization of the collected post-vaccination titers, ignoring all variables except for the assay strain.

Qualitatively summarizing the distribution of titers to all of the assay strains from plots alone is difficult, and the models in the main text are very helpful for understanding the variation in post-vaccination titers. However, we can make a few observations. Most people had some prior immunity (Figure 2) to the A(H3N2) strains which have circulated since the 80’s or 90’s, with protective (40 or greater) titers to the strains from the 2000’s and onward. However, most people only had protective titers to the two most recent A(H1N1) strains, CA/09 and MI/15 which represent the 2009 pandemic lineage. Some people had immunity to older strains, but the difference was much more stark than for A(H3N2). Many people had prior immunity to all of the B strains we examined, and the median was 40 or greater for all of the B strains except MD/59.

Post-titers were, in general, higher (Figure 3). The two pandemic-like A(H1N1) strains showed a boost on average in the population, and there was noticeable back-boosting to some of the older A(H1N1) strains. Many of the A(H3N2) strains showed backboosting as well, although there was not much of a response to the oldest H3N2 strains which also had low pretiters. The median post-titers were above 40 for all of the B strains in our data, with B/Yamagata having the highest average titers, followed by B/Victoria and then the older (B/Pre) lineages.

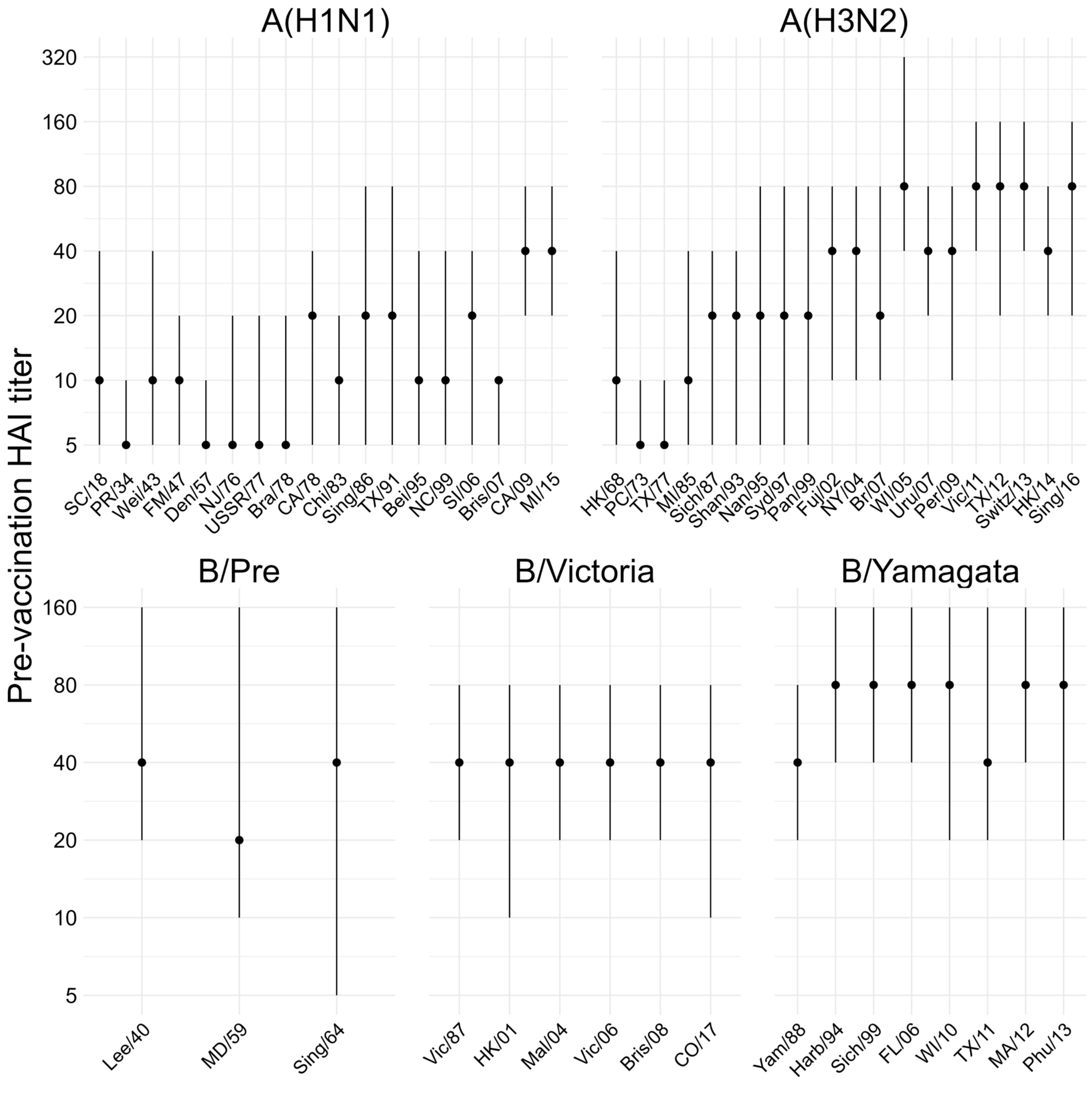

*Figure 2: Pre-vaccination titers in our study to each of the assay strains. The point shows the median and the line shows the IQR.*

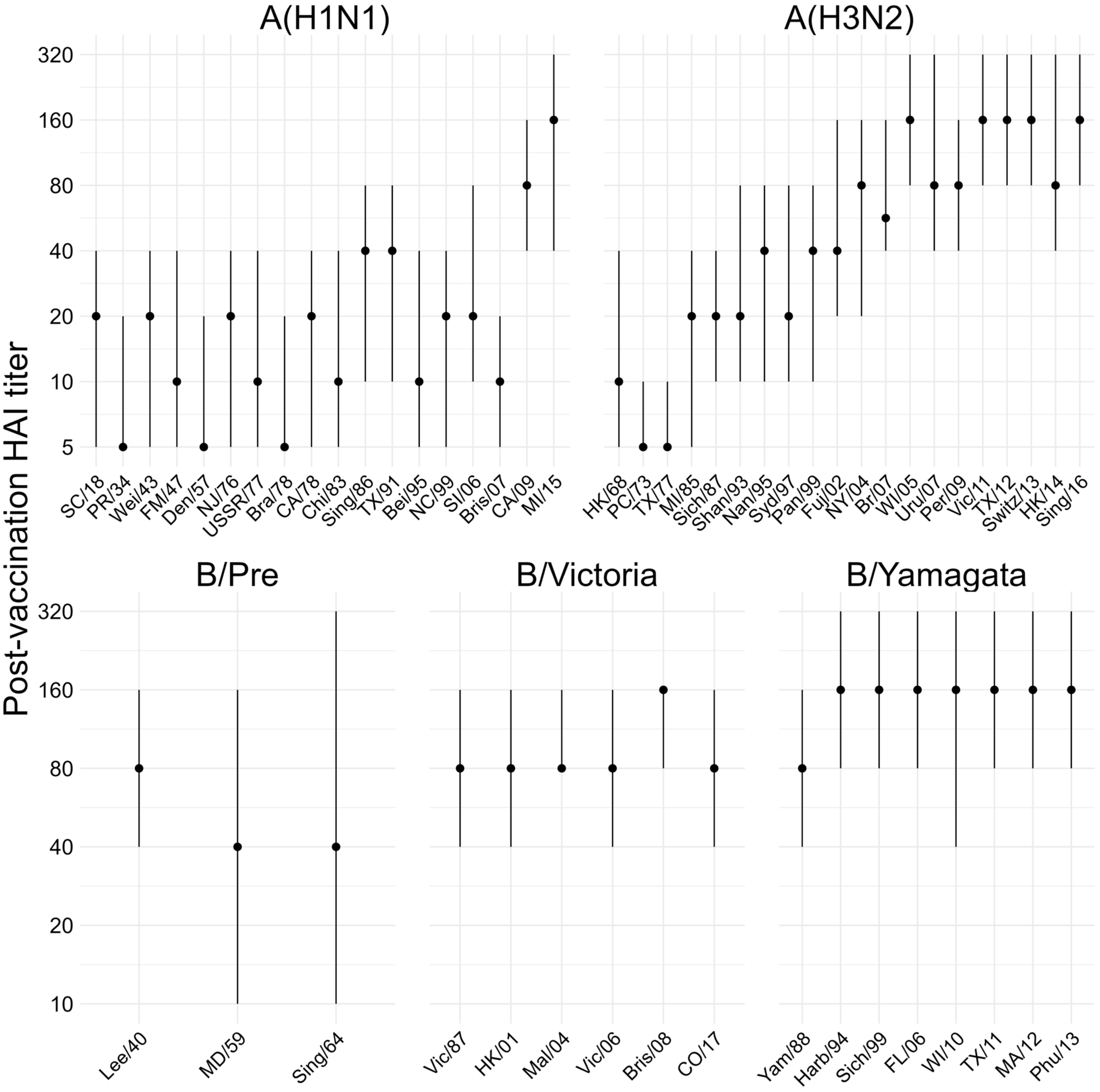

*Figure 3: Post-vaccination titers in our study to each of the assay strains. The point shows the median and the line shows the IQR.*

#### 3.5 Metric agreement analysis

Before we built statistical models for the post-vaccination titer, we first performed a simple unadjusted analysis of the consistency (or agreement) between the antigenic distance measurements. As an omnibus test of agreement, we calculated the intraclass correlation (ICC) across the four antigenic distance measurements, separately for each strain type. We used a Bayesian model with a fixed effect for antigenic distance metric and random intercepts for both assay strain and vaccine strain, and calculated the ICC as the ratio of variance explained by the assay and vaccine strain variance components to the total variation. The Spearman rank correlations show in the main text can be viewed as a post-hoc analysis of the ICC which provide more information about specific comparisons.

Specifically, the model we fit for each subtype can be written as follows.

$$
\begin{aligned}
d[i] &\sim \mathcal{N}(\mu[i], \sigma^2) \\
\mu[i] &= \alpha[1] \cdot I(\text{method}[i] = \text{temporal}) + \alpha[2] \cdot I(\text{method}[i] = \text{p-Epitope}) + \\
&\quad\ \alpha[3] \cdot I(\text{method}[i] = \text{Grantham}) + \alpha[4] \cdot I(\text{method}[i] = \text{cartographic}) + \\
&\quad\ u\big[1, \text{assay strain}[i]\big] + u\big[2, \text{vaccine strain}[i]\big] \\
\alpha[k] &\sim t(3,0,5); \quad k = 1,2,3,4 \\
u[r,\cdot] &\sim \mathcal{N}(0, \zeta[r]); \quad r = 1,2 \\
\zeta[r] &\sim t^+(3,0,1) \\
\sigma &\sim t^+(3,0,1)
\end{aligned}
$$

We fit the model using Stan's NUTS sampler using 12 chains, each with 1000 warmup iterations and 1000 post-warmup sampling iterations and an adaptive delta of 0.99. Model diagnostics were all sufficient (data not shown, the model is easy to sample from and samples quickly). We then calculate the ICC as

$$
\text{ICC} = \frac{\zeta[1]^2 + \zeta[2]^2}{\zeta[1]^2 + \zeta[2]^2 + \sigma^2},
$$

over the posterior samples of all parameters. That is, the ICC represents the ratio of variance due to strain effects only to the total variance after controlling for fixed effects. In the psychometric literature, this is referred to as a one-way ICC for consistency – if the ICC is close to one, it means the variance from the random effects dominates the model. We summarized the ICC as the mean and 95% HDCI across the posterior samples.

As a sensitivity analysis, we considered an alternative agreement statistic based on a different variance decomposition. We fit the same models as before, but then computed the variance of the posterior predictions for every point in the dataset without taking the random effects into account (the "fixed effects" predictions), i.e.

$$
\sigma^2_{\text{FE}} = \text{Var}_{i=1}^{n}\big(\alpha\big[\text{method}[i]\big]\big),
$$

where we choose the correct $\alpha$ parameter based on the method for dataset entry $i$ (we omit writing all four alpha parameters and indicator functions for readability). Then, we compute the variance of the posterior predictions for each entry in the dataset taking the random effects and fixed effects into account:

$$
\sigma^2_{\text{ME}} = \text{Var}_{i=1}^{n}\big(\alpha\big[\text{method}[i]\big] + u\big[1, \text{assay strain}[i]\big] + u\big[2, \text{vaccine strain}[i]\big]\big).
$$

We can then compute an alternative agreement statistic as the variance ratio

$$1 - \sigma^2_{\mathrm{FE}} / \sigma^2_{\mathrm{ME}},$$

which will be close to one if the random effects dominate the prediction variance, or close to zero if the random effects have only a small contribution to the prediction variance. Table 6 shows our results using this metric. All of the results indicate low agreement but with a much higher uncertainty, and this metric is less charitable to the A(H3N2) consistency, although we observed strong pairwise correlations between all of the A(H3N2) metrics as shown in the main text.

*Table 6: Prediction variance ratio across all antigenic distance measurements, calculated separately for each subtype or lineage (strain type). The posterior distribution for each ratio was calculated as one minus the ratio of the prediction variance ignoring random effects to the prediction variance including random effects, estimated with a Bayesian model. Numbers shown are the mean and 95% highest density credible interval (HDCI) of the posterior distribution of variance ratios.*

| Strain Type | PPD Ratio |
|---|---|
| H1N1 | 0.03 (-0.26, 0.32) |
| H3N2 | 0.21 (0.01, 0.39) |
| B-Yam | 0.14 (-0.23, 0.49) |
| B-Vic | -0.04 (-0.75, 0.55) |

#### 3.6 Correlation coefficients and CIs

Table 7 shows the Spearman correlation coefficients and 95% HDCIs for the correlations shown in Figure 1 of the manuscript. The estimates and CIs are arranged by subtype in the table in the same order in which they are shown in the plot. The coefficients may be slightly different from the point estimates we presented in the main text due to rounding error. We calculated the estimates and HDCIs shown here as the mean and 95% HDCI of 4000 samples from a posterior distribution created by Bayesian bootstrapping, which we performed independently on each subtype using 4000 resamples of the observed data points.

Notably, the credible intervals are quite wide for all subtypes other than A(H3N2), which showed strong agreement in both the reliability and correlation assessments. For influenza B subtypes, the width of the credible intervals is almost certainly due to the low number of vaccine/assay strain pairs we observed in our dataset. For A(H1N1), we suspect that the wide credible intervals are due to the multiple clusters in the data, which could violate the assumptions of calculating a correlation coefficient (specifically, Spearman's rank correlation assumes that the rank distributions are bivariate normal between the two variables of interest, which is unlikely to hold in a variable which represents information from multiple heterogeneous clusters). However, our point estimates which reflect low overall agreement are A(H1N1) are similar to the previous results obtained by Bedford et al. (36), and large credible intervals can indicate the lack of a strong signal in the data, so we feel that the conclusions presented in the main text (a lack of agreement across metrics for A(H1N1) and B subtype, and a paucity of influenza B data) are not affected by the presence of wide credible intervals for the correlation coefficients.

*Table 7: Spearman correlation coefficients and 95% HDCIs estimated by Bayesian bootstrap for each influenza subtype. Each pairwise comparison is shown only once to prevent confusion.*

| Subtype | | Cartographic | p-Epitope | Grantham |
|---|---|---|---|---|
| A(H1N1) | Temporal | 0.45 ( 0.13, 0.74) | -0.10 (-0.49, 0.33) | -0.25 (-0.64, 0.17) |
| | Cartographic | | 0.56 ( 0.29, 0.82) | 0.35 (-0.01, 0.68) |
| | p-Epitope | | | 0.90 ( 0.80, 0.97) |
| A(H3N2) | Temporal | 0.93 ( 0.88, 0.97) | 0.88 ( 0.80, 0.95) | 0.96 ( 0.93, 0.98) |
| | Cartographic | | 0.89 ( 0.83, 0.95) | 0.93 ( 0.88, 0.97) |
| | p-Epitope | | | 0.85 ( 0.72, 0.96) |
| B/Yamagata | Temporal | 0.66 ( 0.35, 0.89) | 0.88 ( 0.76, 0.97) | 0.83 ( 0.67, 0.97) |
| | Cartographic | | 0.66 ( 0.39, 0.87) | 0.67 ( 0.36, 0.91) |
| | p-Epitope | | | 0.88 ( 0.73, 0.98) |
| B/Victoria | Temporal | 0.55 ( 0.01, 0.97) | 0.63 ( 0.14, 0.97) | 0.71 ( 0.31, 0.99) |
| | Cartographic | | 0.49 (-0.08, 0.95) | 0.37 (-0.24, 0.88) |
| | p-Epitope | | | 0.92 ( 0.73, 1.00) |
| Overall | Temporal | 0.77 ( 0.67, 0.86) | 0.63 ( 0.48, 0.75) | 0.55 ( 0.39, 0.70) |

| | | | |
|---|---|---|---|
| | Cartographic | 0.82 ( 0.74, 0.89) | 0.77 ( 0.68, 0.86) |
| | p-Epitope | | 0.90 ( 0.85, 0.94) |

#### 3.7 Antigenic distance evenness and dispersion analysis

Since some of the antigenic distance metrics are more discrete than others, we calculated the gap standard deviation as a measure of evenness of distribution across each metric. The gap standard deviation is calculated as the standard deviation of the consecutive differences in the sorted antigenic distance values for a given metric. That is, assume $x$, a vector of measurements from $i = 1, \ldots, n$ is already sorted in increasing order so that $x_1 \leq x_2 \leq \cdots \leq x_n$. Then, the gap standard deviation is computed as

$$\begin{aligned} d_k &= x_{k+1} - x_k; \quad k = 1, \ldots, i-1 \\ \bar{d} &= \frac{1}{n} \sum_{k=1}^{i-1} d_i \\ \sigma_{\text{gap}} &= \sqrt{\frac{1}{n-2} \sum_{k=1}^{i-1} \left(d_k - \bar{d}\right)^2}. \end{aligned}$$

For a random variable with a uniform distribution,

$$\lim_{n \to \infty} \sigma_{\text{gap}} = 0.$$

The different antigenic distance metrics also have different distributions in the set of observed variables. Rather than a uniform distribution of data points across distance space, each metric had gaps in the distribution of observed distances, which varied by metric and subtype (Figure 4 A). The two B lineages had much larger gaps due to the sparser historical panels. For influenza A, all metrics were more uniform for A(H3N2) than for A(H1N1), suggesting their different evolutionary patterns across the time spanned by the historical panel. Notably, while the temporal metric was the most uniform for all strains (an artifact of how the historical panel was chosen), the Grantham and $p$-Epitope metrics tend to discretize the number of potential distances and result in less uniformly distributed values for the historical panel used in our study.

We quantified the uniform spread of points for each antigenic distance metric and subtype using the gap standard deviation, where a higher gap standard deviation indicates more irregularity in the spacing of data points. Figure 4 B shows the estimated gap standard deviations. Both B lineages had higher gap standard deviations for all methods than either influenza A subtype. For A(H3N2), the gap standard deviations were similar across antigenic distance methods, and for A(H1N1) the differences were still small but larger than A(H3N2), representing the diversity of strains in the historical panel for type A influenza strains. The differences were much more noticeable for both B lineages, with Grantham distance having notably higher gap standard deviation than the other metrics for both influenza B lineages, indicating lower diversity in the normalized distance values.

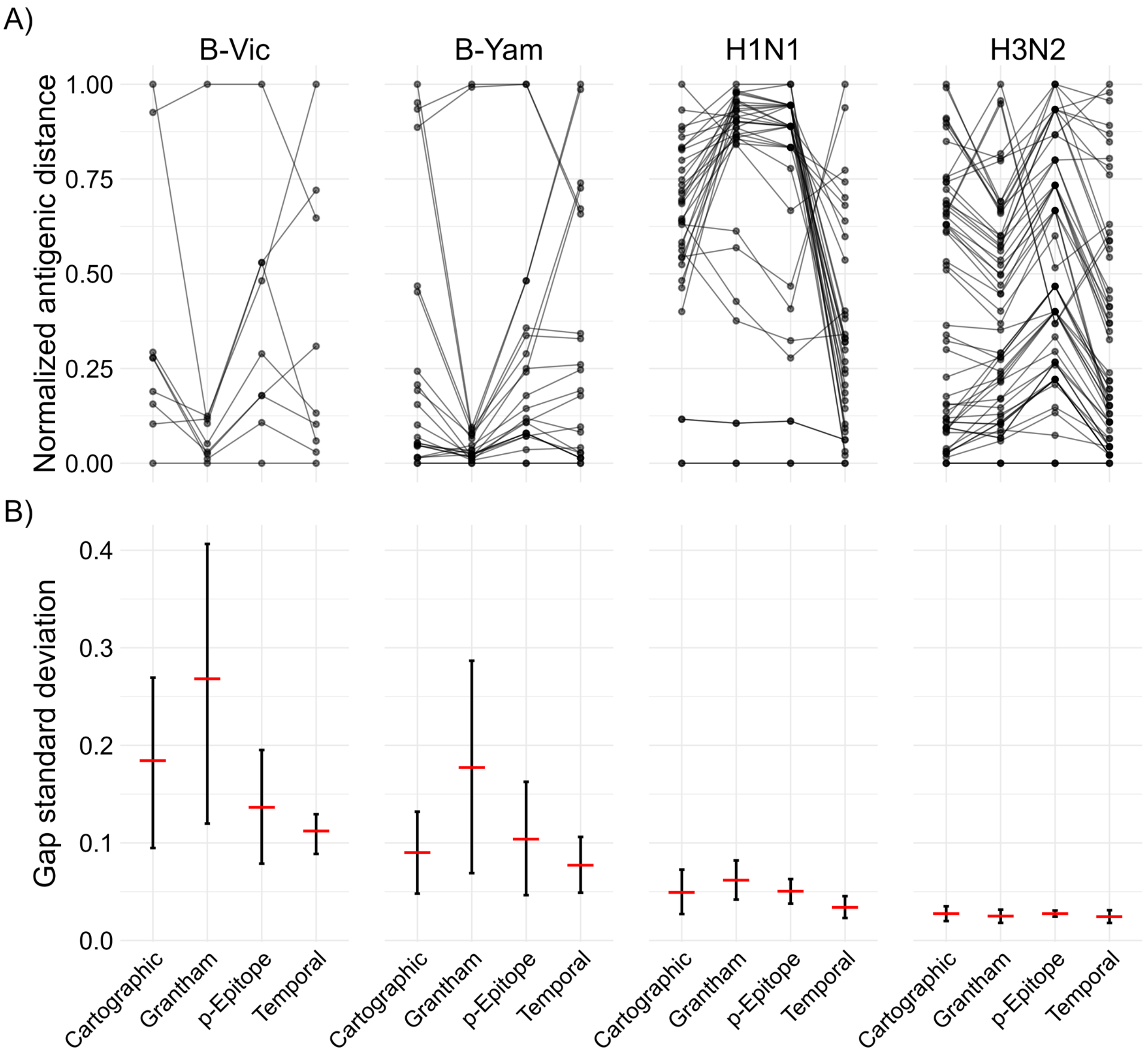

*Figure 4: A) Parallel coordinates plot showing how the estimated pairwise antigenic distances change for each of the antigenic distance metrics. Each line in the plot represents one vaccine strain and assay strain pair, and the connected points are the pairwise distance measured under each metric shown on the x-axis. When two lines cross, this indicates that two metrics assigned a different relative order to the pairwise combination. Note that Grantham and especially $p$-Epitope distances are integer-valued and concentrate measurements to specific points which potentially overlap (temporal distance is also integer valued but has enough spread to avoid a similar issue). B) The gap standard deviation (gap SD) for each subtype and antigenic distance metric. The posterior distribution of gap SDs was calculated using the bayesian bootstrap with*

*reweighting. The red horizontal bar shows the mean of the bootstrap posterior and the error bars show the 95% highest density credible interval (HDCI).*

#### 3.8 Model diagnostics

We examined the key model diagnostics for all of our models to ensure they converged. The main diagnostics with target criteria identified by the Stan development team (29) are:

- $\hat{R}$, which measures chain mixing, should be $< 1.01$ for all parameters;
- Bulk and tail ESS, measures of the number of samples drawn if all of the samples were independent, should be greater than 1000;
- Number of divergent transitions should be less than 1% of samples;
- Number of treedepth exceedences should be less than 1% of samples;
- E-BFMI should be greater than 0.3 for all chains.

These diagnostics are presented in Table 8.

*Table 8: Model diagnostics for the GAMMs and LMMs fit with each of the antigenic distance metrics. We show the total number of divergences out of the number of samples along with other common diagnostic criteria. For each model, we show the minimum ESS across all parameters, the minimum E-BFMI across chains, and the maximum R hat across all parameters.*

| Model | Pct. Divergences | min ESS (tail) | min ESS (bulk) | min E-BFMI | max R_hat |
|---|---|---|---|---|---|
| GAMM | 0.0% | 5593 | 2574 | 0.48 | 1.02 |
| | 0.2% | 6128 | 2831 | 0.51 | 1.02 |
| | 0.0% | 2820 | 1483 | 0.45 | 1.03 |
| | 0.0% | 4265 | 2103 | 0.52 | 1.02 |
| | 0.1% | 3887 | 1914 | 0.52 | 1.02 |
| | 0.0% | 3953 | 1870 | 0.48 | 1.02 |
| LMM | 0.0% | 4293 | 2608 | 0.46 | 1.02 |
| | 0.0% | 4325 | 2430 | 0.53 | 1.02 |
| | 0.3% | 4286 | 1923 | 0.50 | 1.02 |
| | 0.0% | 3967 | 1554 | 0.50 | 1.02 |
| | 0.0% | 3903 | 2290 | 0.49 | 1.02 |
| | 0.0% | 4953 | 2398 | 0.52 | 1.02 |

Most of our models had $\hat{R}$ statistics which were 1.02 or 1.03, but these were for highly constrained parameters. Each model already takes at least 6 days to run in an HPC environment, so running the models longer is computationally infeasible due to the size of our dataset and unlikely to qualitatively change our results.

We also examined trace plots of the parameters to ensure there were no obvious errors (and, in general, errors in the trace plots will be noticeable in the $\hat{R}$ statistic). We also examined the prior/posterior shrinkage and visually inspected prior/posterior plots. Since we have many models, each with thousands of parameters, we did not include the plots here. We observed good values of shrinkage (far from 1, indicating a divergence away from the prior) for most parameters, with the exception of some highly constrained parameters, typically correlations and GAMM regularizing variance parameters. Some of the random effects for individuals had poor shrinkage as well, but overall the shrinkage for random effects and for the random effects variances was far from 1. Since the GAMM was not supported by ELPD anyways, we did not investigate prior sensitivity analysis further since all of the LMM parameters had good shrinkage. Therefore, we feel safe about our choice of regularizing priors and a prior sensitivity analysis would require extensive computational time without being useful.

#### 3.9 Prior sampling diagnostics

While less important for our purposes, we also sampled from the priors in order to examine the prior/posterior shrinkage and to visualize our prior predictive simulations. Such an analysis requires substantially less computational power than sampling from the posterior distribution, but we still need to ensure that we have sampled from the priors enough to get good estimates of the prior distributions of some highly constrained parameters. So, our prior sampling diagnostics are shown in Table 9.

*Table 9: Model diagnostics for samples from the prior distributions for our GAMMs and LMMs. These samples are drawn only from the prior distributions and do not see the data. For each model, we show the minimum ESS across all parameters, the minimum E-BFMI across chains, and the maximum R hat across all parameters.*

| Model | Pct. Divergences | min ESS (tail) | min ESS (bulk) | min E-BFMI | max R_hat |
|---|---|---|---|---|---|
| GAMM | 0.0% | 7920 | 7008 | 0.82 | 1.01 |
| | 0.0% | 9218 | 7041 | 0.84 | 1.01 |
| | 0.0% | 8789 | 7519 | 0.85 | 1.01 |
| | 0.0% | 9100 | 7097 | 0.84 | 1.01 |
| | 0.0% | 9013 | 6603 | 0.87 | 1.01 |
| | 0.0% | 8805 | 7179 | 0.78 | 1.01 |
| LMM | 0.0% | 7743 | 6715 | 0.85 | 1.01 |

| | | | | |
|---|---|---|---|---|
| 0.0% | 7412 | 6429 | 0.82 | 1.01 |
| 0.0% | 6843 | 6583 | 0.86 | 1.01 |
| 0.0% | 7793 | 6664 | 0.86 | 1.01 |
| 0.0% | 7794 | 6586 | 0.81 | 1.01 |
| 0.0% | 7618 | 6649 | 0.84 | 1.01 |

Since there are thousands of parameters per model, we do not show the shrinkage parameters or prior distributions of all parameters here, but they are easy to produce from the code and results we provide.

#### 3.10 ELPD Diagnostics

Similar to frequentist AIC/BIC and Bayesian WAIC, ELPD relies on a computationally efficient approximation to leave-one-out cross validation that allows estimation of a goodness-of-fit metric without having to refit a computationally impossible number of models. However, unlike other informatic criteria, the LOO-IC based on the leave-one-out expected log pointwise predictive density provides diagnostics to determine if the approximation is trustworthy (37,38). Table 10 shows the diagnostic measures for each of our models. The maximum Pareto $k$ diagnostic is the primary value indicating whether the LOO-ELPD approximation is accurate – all Pareto $k$ values (one per observation) should be below $0.7$. The $N_{\text{eff}}$ value is the effective sample size for the approximation, and the ratio of the effective sample size to the actual sample size should be greater than $0.5$ to ensure that the threshold of $0.7$ is reliable. If the number of effective samples is greater than 2200 however, the threshold of $0.7$ is useful regardless of the ratio.

*Table 10: Diagnostics for the LOO-IC ELPD approximation. Pareto $k$ is the primary diagnostic indicating whether the approximation is trustworthy and all Pareto $k$ values should be below 0.7. The $N_{eff}$ is the effective sample size and $R_{eff}$ is the ratio of the effective sample size to the true sample size – if there are too few effective samples relative to actual samples, we can get an optimistic evaluation of the approximation quality, but in general this matters less if the ESS is sufficiently high.*

| Metric | Model | Max. Pareto k | Min. N_eff | Max. R_eff |
|---|---|---|---|---|
| Cartographic | GAMM | 0.30 | 4429.2 | 1.00 |
| | LMM | 0.39 | 3522.5 | 1.00 |
| p-Epitope | GAMM | 0.37 | 4179.3 | 1.00 |
| | LMM | 0.37 | 4355.5 | 1.00 |
| Grantham | GAMM | 0.36 | 3883.5 | 1.00 |

| | | | | |
|---|---|---|---|---|
| | LMM | 0.41 | 3547.4 | 1.00 |
| Temporal | GAMM | 0.37 | 3788.6 | 1.00 |
| | LMM | 0.38 | 3525.1 | 1.00 |

#### 3.11 Pointwise prediction comparisons

To examine the difference in predictions across each of the antigenic distance metrics, we computed the fold change in predicted post-vaccination HAI titer conditional on normalized antigenic distance and strain type for each unique pair of antigenic distance metrics. We visually inspected the conditional fold changes between metrics using a limit of agreement approach with a clinically defined threshold for whether the difference between predictions should matter, which is commonly defined as a 4-fold change for HAI measurements. We performed this fold change between predictions analysis for both the GAMM and LMM with each antigenic distance metric.

Figure 5 shows the prediction comparisons across antigenic distance metrics for each subtype using the LMMs. In contrast to our agreement analysis, where the A(H3N2) metrics showed the strongest agreement across metrics (and the highest pairwise correlations), A(H3N2) was the only subtype with noticeable trends in the contrasts between metrics. In particular, all of the comparisons with $p$-Epitope for A(H3N2) had a noticeable trend – even though the mean fold change in predictions always stayed within the measurement error boundaries we set *a priori*, sometimes the credible interval did not fully cover the measurement error boundaries and there was a noticeable slope. These trends suggested that $p$-Epitope measurements underestimated the expected change in post-vaccination titer compared to Grantham and cartographic distance, while $p$-Epitope overestimated the difference compared to temporal methods. These results suggest that perhaps biochemical features like glycosylation sites or changes to the virus outside of the immunodominant epitope region are important, because these features are detected by cartographic and Grantham distance, but not by $p$-Epitope distance.

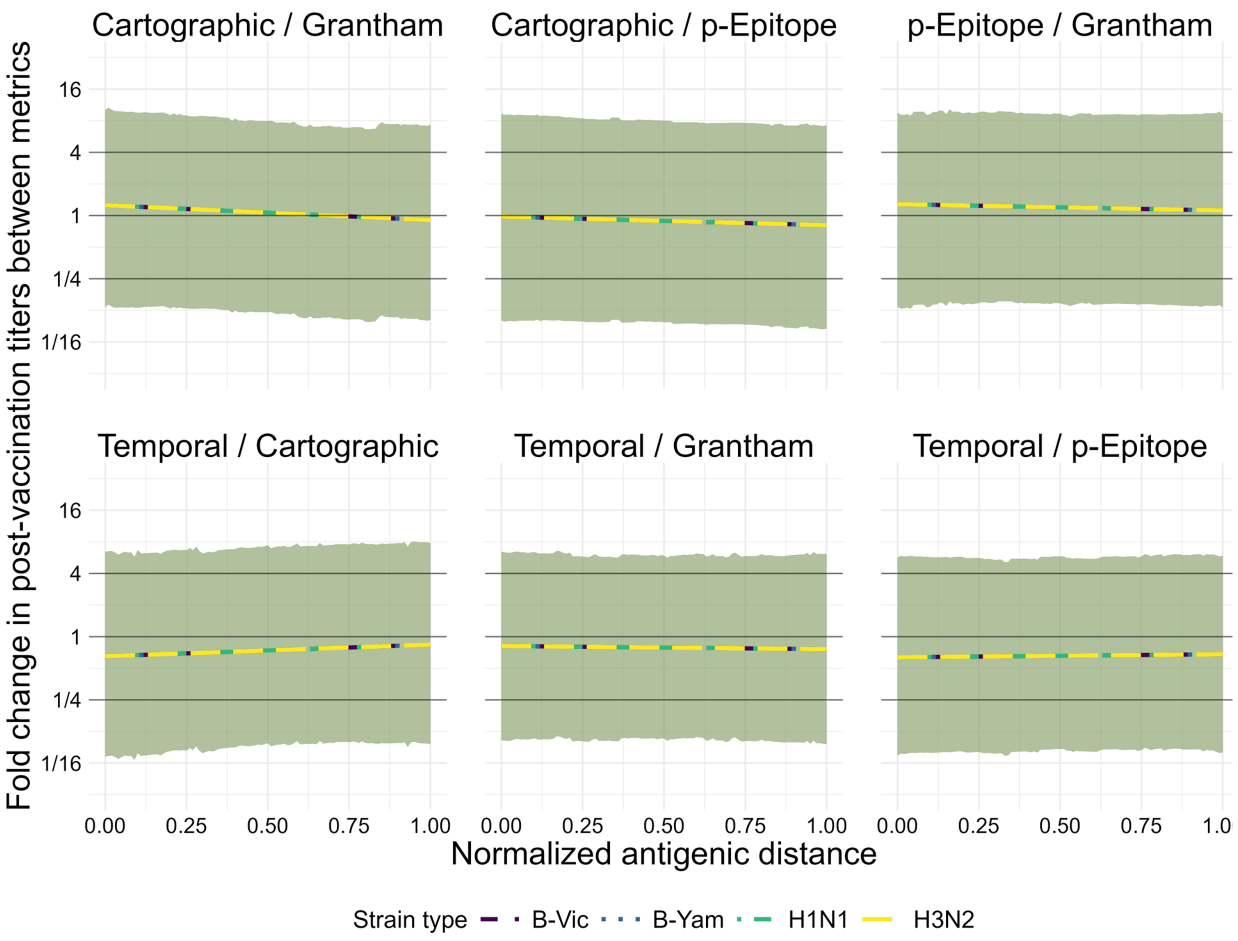

*Figure 5: Pairwise comparisons of predictions (from the LMMs) between each unique set of two metrics. The y-axis shows the fold change in predictive titers between metrics, and the two metrics being compared in each subplot are shown as the subplot labels. Each line represents the predictions for the first metric in the pair at a given antigenic distance value divided by the predictions for the second metric in the pair. Color and linetype correspond to different strain types. The solid black lines on the plot are reference lines at a value of 1 for no effect, and at 4 and 1/4, effect values which would represent a clinically notable deviation in HAI predictions beyond what is expected from measurement error. Lines represent the mean of the posterior distribution of the contrast and the colored ribbons represent the 95% highest density credible interval (HDCI) for each strain type in each subplot.*

Figure 6 shows the prediction comparisons across antigenic distance metrics for each subtype using the GAMMs. Even though the GAMM was not supported by our ELPD analysis, we used the GAMM for analyzing pairwise differences in predictions in case the nonlinear signal was biologically important with a weak signal. Unlike our simple correlation analysis, this analysis examines the predicted protection for an average individual exposed to an antigenically distant strain after vaccination, rather than only taking antigenic distance into account. We saw that the fold change in predicted HAI titers was almost always less than four for every pairwise comparison between two metrics. A four-fold change in HAI titer is considered a clinically relevant difference between two measurements, so in almost every case we saw that changing the antigenic distance metric would not lead to a clinically relevant difference in predicted post-vaccination HAI titer. The primary exception was strain type A(H1N1), which exceed 40 at a few antigenic distance values for some of the pairwise comparisons (around a normalized antigenic distance of 0.25 for the cartographic/Grantham and Cartographic/p-Epitope comparisons, and around a normalized antigenic distance of 0.75 for the Grantham/temporal distance comparisons). Due to the large standard errors and the number of comparisons we make, we are comfortable attributing these fluctuations to measurement error, although the large variability across antigenic clusters for A(H1N1) strains (pdm-like vs. non-pdm-like) could contribute as well.

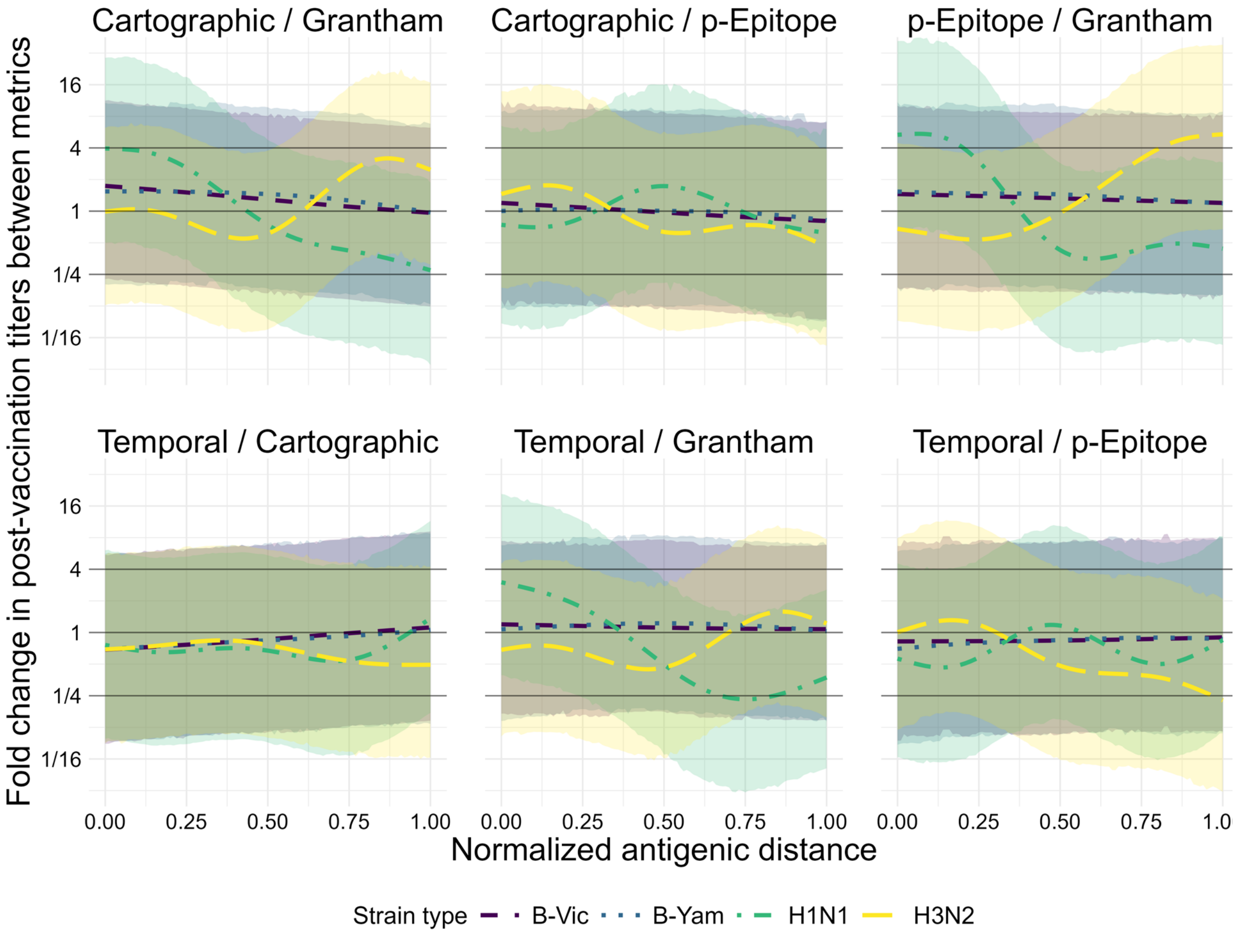

*Figure 6: Pairwise comparisons of predictions (from the GAMMs) between each unique set of two metrics. The y-axis shows the fold change in predictive titers between metrics, and the two metrics being compared in each subplot are shown as the subplot labels. Each line represents the predictions for the first metric in the pair at a given antigenic distance value divided by the predictions for the second metric in the pair. Color and*

*linetype correspond to different strain types. The solid black lines on the plot are reference lines at a value of 1 for no effect, and at 4 and 1/4, effect values which would represent a clinically notable deviation in HAI predictions beyond what is expected from measurement error. Lines represent the mean of the posterior distribution of the contrast and the colored ribbons represent the 95% highest density credible interval (HDCI) for each strain type in each subplot.*

However, the differences in comparisons for A(H3N2) was not completely trivial either. Figure 6 shows that for A(H3N2), the temporal distance overwhelming underestimates the fold change in predictions for the largest antigenic distances compared to both Grantham and $p$-Epitope measurements, with some interesting trends in the comparisons between cartographic distance as well. These results support our conclusion that further research into which of these metrics actually captures useful and interesting features is warranted, because it is difficult to tell whether we are capturing noise from our study or actual patterns that suggest different metrics are identifying different relevant characteristics of the viruses.

In both models, nearly all contrast predictions fall within the clinically irrelevant reference bounds, although the credible intervals for all predictions are wide because our bayesian models fairly account for many sources of uncertainty in the data. However, our results for the GAMM model suggest some interesting exceptions for the A(H1N1) strains that are likely related to the pandemic-like and non-pandemic-like cluster differences. Our results for the GAMM and LMM model for A(H3N2) seem to suggest that perhaps different metrics are picking up different relevant features, as we noted in the main text discussion.

#### 3.12 Vaccine-specific Predictions

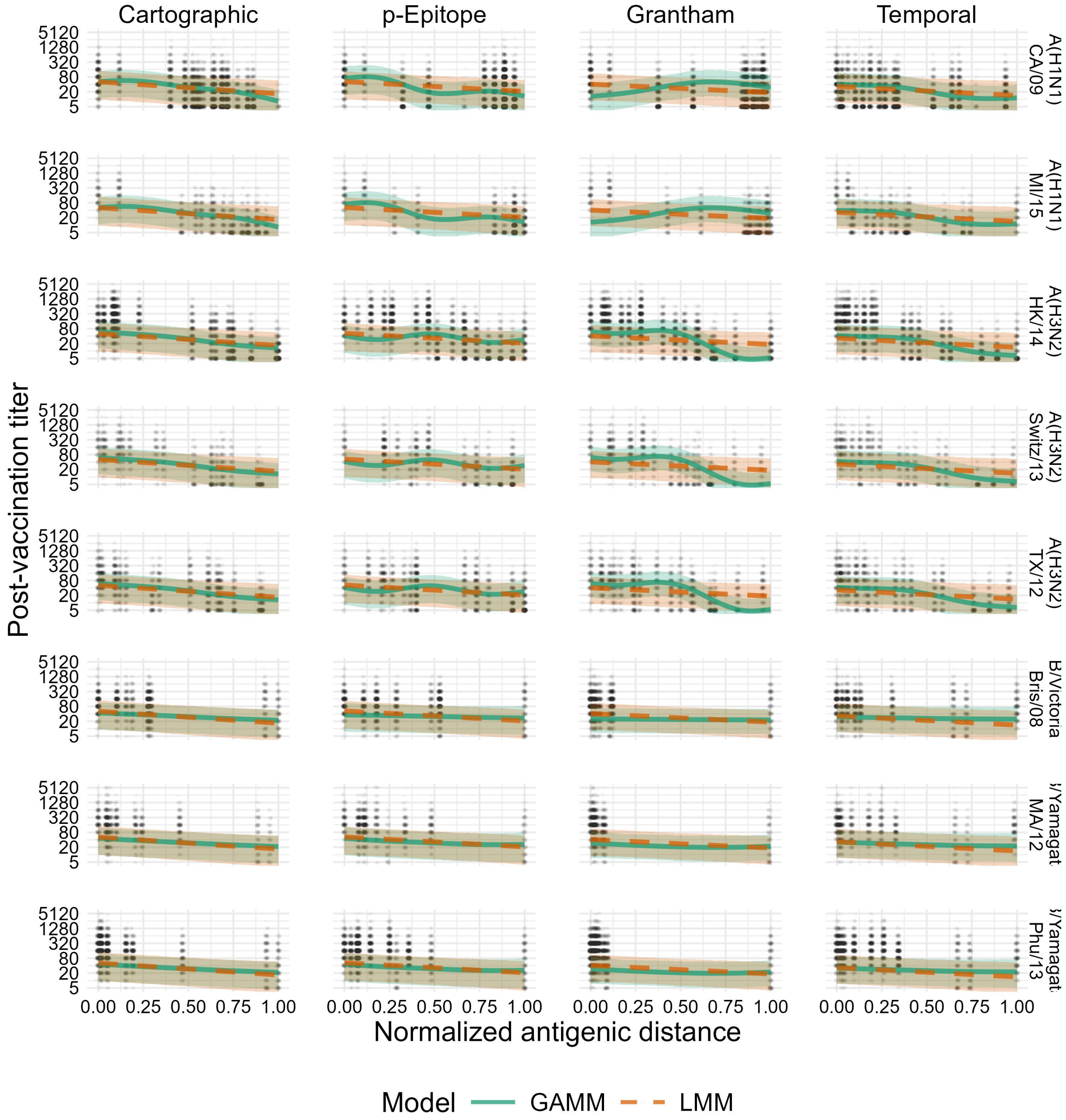

*Figure 7: Model predictions for both the GAMM and LMM, conditional on the vaccine strain rather than only on the subtype (shown in the main text). Solid green lines and green ribbons show the mean and 95% highest density continuous interval (HDCI) for GAMM predictions. Dashed orange lines and orange ribbons show the mean and 95% HDCI for LMM predictions. Circular points show the data values. Each subplot shows the model predictions for a particular subtype (changes by row) and distance metric*

*(changes by column). Outcomes shown on the plot are predicted post-vaccination titers for an average individual to an average strain.*

In order to analyze the differences between vaccine strains, we also examined the results conditional on the specific strain used in a vaccine (for a given subtype). Within a given subtype, there were no striking results across the different vaccine components. In the main text, we show that direct causal effects of vaccine and assay strain contribute very little to the variance in the outcomes after controlling for antigenic distance.

#### 3.13 Alternative distance metrics

Because we chose to include the dominant $p$-Epitope distance and Grantham's distance in our final manuscript, we also analyzed other sequence-based and biochemical distances to determine if our arbitrary choice was misleading and we should consider further antigenic distance measures. So, we compared the dominant $p$-Epitope distance (2,39) with the $p$-all-Epitope distance (39,40) and the Hamming distance (41). Furthermore, we compared Grantham's distance (4) with the Hamming distance and with the FLU substitution model, an evolutionary amino acid substitution matrix model derived specifically for influenza sequences (42) (there are other indices like Grantham's index, but we felt that the comparison to a model specifically for influenza amino acid substitutions was sufficient).

When we compared the FLU substitution model to Grantham's distance and the simple Hamming distance, we found that all three metrics were highly correlated for all subtypes except A(H1N1), with relatively small credible intervals from Bayesian bootstrapping (Table 11). For A(H1N1), Grantham and Hamming distances were also highly correlated, but the correlations between Hamming and FLU substitution and Grantham and FLU substitution distances were moderate at best, with credible intervals that covered quite low values. The original study which developed the FLU substitution matrix used a mix of influenza virus sequences across multiple proteins and types/subtypes (42), so it is unclear why the difference would be so stark for A(H1N1). Regardless, because the difference was only noticeable for A(H1N1) we decided to use the Grantham distance in our main analysis. Despite high similarity to the Hamming distance across all subtypes, Grantham distance contains more information by design and better antigenic coverage of Influenza B strains in a future study might reveal further differences between Grantham distance and Hamming distance.

*Table 11: Pairwise Spearman rank correlations between antigenic distance values using the Grantham, FLU Substitution, and Hamming distance metrics. We calculated correlations between two distances using the normalized distance values between every vaccine/assay strain pair for the given subtype. Numbers shown are the mean and 95% highest density continuous interval (HDCI) calculated by Bayesian bootstrapping.*

| Subtype | | Grantham | FLU Substitution |
|---|---|---|---|
| A(H1N1) | Hamming | 0.91 ( 0.82, 0.98) | 0.62 ( 0.35, 0.85) |
| | Grantham | | 0.41 ( 0.05, 0.75) |
| A(H3N2) | Hamming | 0.99 ( 0.98, 1.00) | 0.97 ( 0.94, 0.99) |
| | Grantham | | 0.96 ( 0.93, 0.98) |
| B/Yamagata | Hamming | 0.98 ( 0.93, 1.00) | 0.94 ( 0.86, 1.00) |
| | Grantham | | 0.91 ( 0.80, 0.98) |
| B/Victoria | Hamming | 0.99 ( 0.97, 1.00) | 0.97 ( 0.89, 1.00) |
| | Grantham | | 0.97 ( 0.88, 1.00) |
| Overall | Hamming | 0.99 ( 0.97, 1.00) | 0.94 ( 0.91, 0.96) |
| | Grantham | | 0.91 ( 0.88, 0.95) |

We also examined the pairwise Spearman correlations between the Hamming distance, the (dominant) $p$-Epitope method which we present in the main analysis, and the $p$-all-Epitope distance, which is calculated by averaging the Hamming distance across all 5 of the immunodominant HA epitope sites. Again, the correlations were overall high Table 12, with A(H1N1) displaying a noteably lower correlation across differences. These supplementary results suggest that different antigenic distance metrics may have the strongest effect on understanding the immune response to A(H1N1), probably in accounting for noteable differences across clusters. The multiple clusters in A(H1N1) antigens are the primary differentiating factor from the ladder-like continuously evolutionary pattern in A(H3N2) and might explain the differences, although we lack the ability to analyze this further.

*Table 12: Pairwise Spearman rank correlations between antigenic distance values using the Grantham, FLU Substitution, and Hamming distance metrics. We calculated correlations between two distances using the normalized distance values between every vaccine/assay strain pair for the given subtype. Numbers shown are the mean and 95% highest density continuous interval (HDCI) calculated by Bayesian bootstrapping.*

| Subtype | | p-Epitope | p-All-Epitope |
|---|---|---|---|
| A(H1N1) | Hamming | 0.93 ( 0.87, 0.98) | 0.89 ( 0.79, 0.98) |
| | p-Epitope | | 0.89 ( 0.79, 0.97) |
| A(H3N2) | Hamming | 0.85 ( 0.71, 0.97) | 0.92 ( 0.83, 0.99) |

| | | | |
|---|---|---|---|
| | p-Epitope | | 0.97 ( 0.94, 0.99) |
| B/Yamagata | Hamming | 0.92 ( 0.82, 0.99) | 0.96 ( 0.90, 0.99) |
| | p-Epitope | | 0.97 ( 0.91, 1.00) |
| B/Victoria | Hamming | 0.94 ( 0.80, 1.00) | 0.99 ( 0.97, 1.00) |
| | p-Epitope | | 0.95 ( 0.84, 1.00) |
| Overall | Hamming | 0.91 ( 0.86, 0.95) | 0.95 ( 0.92, 0.98) |
| | p-Epitope | | 0.97 ( 0.95, 0.98) |

Due to the relative consistency across these other antigenic distance metrics, we did not fit further models to other antigenic distance metrics. The models require a great deal of computational time and power, and since we found overall good agreement between these additional metrics (and a large amount of disagreement within A(H1N1), as we found for the metrics in the main analysis), we felt that this did not justify a further investigation. However, a future study with an expansive panel of serological data to A(H1N1) and A(H3N2) strains to further explore why A(H1N1) metrics have lower agreement would be useful for further understanding these results.
